# Digital genetic counselling services for cascade cardiogenetic testing: a focus group study on proband, relative, and provider perspectives

**DOI:** 10.1101/2024.11.27.24318108

**Authors:** Marlies N. van Lingen, Sietske A. L. van Till, Noor A.A. Giesbertz, Tessa C. Beinema, Margreet G.E.M. Ausems, Randy Klaassen, Martina C. Cornel, Lieke M. van den Heuvel, J.Peter van Tintelen

## Abstract

Digital interventions are potentially promising to improve accessibility and efficiency of genetic counselling services. However, current literature on stakeholder perspectives towards digital tools for cascade testing is limited. Therefore, this focus group study aimed to gain insights into the attitude and perspectives of probands, at-risk relatives (ARR), and genetic healthcare professionals (HCP) towards digital innovations for assistance with both pre-test and post-test counselling and cascade genetic testing in cardiogenetics. We conducted seven online focus groups, which where transcribed and thematically analysed. In total, 37 individuals participated (10 probands, 11 ARR and 16 HCP). Thematic analysis of focus group transcripts showed a first theme of (1) acceptability of digital tools. Other identified themes were defined as ‘domains’ where digital tools impact traditional, in-person clinical genetic care, being: (2) family communication, (3) decision-making, (4) care relations, and (5) the genetic care system. Stakeholders expressed a predominantly positive attitude towards digitisation of (parts of) the predictive genetic counselling in cardiogenetics, under the condition that access to human contact is preserved. In the clinical setting of predictive counselling, efforts should be made to ensure access to genetic services for all ARR and to protect in-person involvement of HCP.

## Introduction

The importance of cascade genetic testing for monogenic treatable inheritable diseases is undisputed [1–3]. After identification of the disease-causing variant in a proband, cascade screening in at-risk relatives (ARR) is recommended [4, 5]. In the Netherlands, the family-mediated approach is adopted, where probands inform their ARR about an inherited disease and genetic testing options, assisted by a family letter from their genetic healthcare professional (HCP) [6]. Even though cascade testing, followed by clinical follow-up and treatment when indicated, can decrease morbidity and mortality in ARR [4, 5], the uptake of genetic counselling and/or predictive genetic testing for monogenic treatable diseases ranges between 40% and 60% in cardiogenetics [1, 7–10] and oncogenetics [11–13].

Several barriers have been identified that hamper probands from informing their ARR or that prevent ARR from obtaining genetic counselling, including family-related [2, 14–16], knowledge [15, 16] and practical barriers, including financial barriers, limited access to genetic services [2], or fear of genetic discrimination by insurers [17]. Digital interventions have been identified as potentially promising to improve the accessibility and efficiency of genetic counselling services [18, 19].

Existing digital genetic counselling tools include conversational agents (CA, i.e., chatbots or virtual assistants) [20], decision aids [21], family outreach tools [22], and full pre-[23] and post-test counselling platforms [24]. Development and implementation of digital tools for predictive genetic counselling raises ethical and practical questions, including questions related to equitable access, clinical validity and utility, and involving end-users during the design [18]. As probands, ARR, and HCP are the main actors in genetic counselling, their perspectives towards the development of digital tools are crucial for responsible integration of technologies into genetic care. However, studies on stakeholder perspectives towards digital tools for cascade testing are limited, except for studies on attitudes of patients towards chatbots [25, 26].

A previous focus group study with 29 patients receiving clinically actionable variants showed that most participants were willing to use a follow-up and cascade chatbot while several were willing to share genetic results with ARR via a chatbot [25]. Still, participants expressed privacy and usability concerns in family communication [25]. However, it remains unclear how patients with diverse backgrounds or HCP view CA or other digital technologies for family communication and cascade testing. Moreover, no studies on stakeholder experiences targeting inherited cardiac diseases have been published [19], which are particularly promising for digitalisation because of their prevalence and potential for standardised information provision in predictive counselling. Therefore, this focus group study aims to gain insights into attitudes and perspectives of probands, ARR, and HCP towards digital counselling tools to assist family communication and both pre- and post-test counselling and cascade genetic testing of inherited cardiac diseases.

## Materials and methods

### Design

A qualitative focus group design was chosen to gain insights into the attitude and perspectives of stakeholders and to enhance discussion and forming of new ideas on this novel topic. This study was a preparation for the design and development of “DNA-poli”, an online platform for predictive counselling in cardiogenetic cascade testing. The consolidated criteria for reporting qualitative research (COREQ) were used to report methods and results [27].

### Participants and procedure

Participants were divided into three groups: probands, ARR, and genetic HCP (i.e., clinical geneticists (in training), genetic counsellors, genetic social workers, or genetic laboratory specialists). Probands and ARR were 18 years or over, and a (likely) pathogenic variant in an established gene underlying the proband’s phenotype had to be present. ARR were defined as first- or second-degree family members. Probands and ARR were invited by a letter if they received in-person genetic counselling between October 2020 and October 2021 at the UMC Utrecht. Eligible probands and ARR were invited by four HCP based on inclusion criteria. Genetic HCP were eligible if they were involved in cardiogenetic cascade testing in three Dutch University Medical Centres (Utrecht, Amsterdam, Groningen). Written informed consent was obtained from all participants. After obtaining informed consent, participants were invited via e-mail to an online focus group session (MS Teams, Microsoft, Washington, USA). During each focus group, one researcher (MH) was available to support participants by telephone to address technical questions or problems. After finalising all focus groups, participating probands and ARR were asked to invite their ARR who did not obtain genetic counselling to participate in this study during a follow-up focus group session.

### Data collection

Seven semi-structured focus groups were organised between November 2021 and February 2022. They were scheduled when five to eight participants were available. Probands, ARR, and HCP were clustered in separate focus group sessions. Participants of each stakeholder subtype were actively selected based on age, gender, and digital skills in ARR and probands, and profession for HCP. Participant characteristics were collected prior to the focus group during a telephone consultation by the executing researcher (MvL). These included demographic information (i.e., age, sex, and educational level), information on digital literacy (i.e., ability to perform digital tasks) and health literacy (i.e., ability to read and interpret and health-related information), data related to the cardiogenetic disease (i.e., disease type, relation to proband, onset of disease, carrier status), and information on the method used to inform ARR (Supplement S1).

The semi-structured topic list covered general perspectives on the concept of a digital genetic service, the role of digital technology in informing ARR about genetic risk, online information provision and genetic counselling, role of CA (3 focus groups discussed chatbots and 4 virtual assistants), and considerations regarding decision-making and genetic testing (Supplement S2). HCP were additionally asked about the process of informing probands and organising clinical follow-up in a digital context.

All focus groups were moderated by a trained executing researcher (MvL). Two observers (alternating: ID, MH, LvH, NG, SvT) assisted the moderator with taking field notes and covering the topic list. Sessions were video-recorded (MS Teams) and audio-recorded (Audacity, Version 3.1). Participants received a 10 Euro gift card for an online retailer.

### Data analysis

Audio files were transcribed verbatim. An inductive method of data-generated open coding and attribute coding was independently adopted by two researchers (SvT, MvL), using Nvivo 12 software (Lumivero, Denver, CO, USA). Subsequently, open coding of one focus group of every subgroup (proband/ARR/HCP) was performed, resulting in a draft codebook, after which intercoder differences in codes and definitions were resolved by discussion. After open coding of all focus groups, a final codebook was obtained by a cross-case analysis of the concepts in the original data and the codebook (based on the QUAGOL method [28]). LvdH critically assessed samples of coded transcripts. Based on the codebook, themes and subthemes were defined by consensus between MvL, SvT and LvdH. We reached theoretical saturation of attitudes and perspectives in different groups: only a few new codes were added during final coding of the transcripts and there was consensus about the meaning of the (sub)themes. Illustrating quotes were selected by MvL. Member checking of results was not performed.

## Results

### Study participants

Thirty probands, 46 ARR from 37 families (3 ARR were related a proband), and 30 HCP were invited to participate. No ARR who did not receive genetic counselling responded after being approached by their relatives. Thirteen probands, 13 ARR and 18 HCP (43%, 28%, 60% respectively) consented to participate. Three participants later declined to participate due to personal circumstances. Four participants (one proband, one ARR, two HCP) were unavailable at the actual focus group meeting. In total, 37 individuals participated in seven focus groups (10 probands, 11 ARR, and 16 HCP). Median length of the focus groups was 86 minutes (range 73-104).

Participant characteristics are presented in Table 1. Six out of 11 ARR were first-degree relatives. ARR were directly informed by the proband, and all had received a family letter written by the genetic HCP through the proband. All ARR pursued predictive genetic testing; three ARR (27%) were identified as carriers. Among HCP, clinical geneticist was the most common profession (44%). All participants were born in the Netherlands.

**Table 1.** Participant characteristics. ARR= at-risk relative, HCP=healthcare professional, ACM=arrhythmogenic cardiomyopathy, DCM=dilated cardiomyopathy, HCM=hypertrophic cardiomyopathy. ^1^In probands, clinical phenotype is reported. In at-risk relatives, the phenotype of the affected proband in their family is reported. ^2^Health literacy was defined as medium if participants needed occasional assistance with interpreting and reading health related information from their GP or hospital and low if help was always required. Similarly, digital literacy was understood as needing occasional or constant assistance with digital tasks such as using a computer, visiting a website or using two-factor authentication, represented by medium or low levels of digital literacy, respectively.

| PARTICIPANT CHARACTERISTICS | PROBANDS<br>(N=10) | ARR<br>(N=11) | GENETIC HCP<br>(N=16) |
| --- | --- | --- | --- |
| Sex |  |  |  |
| Female | 6 (60%) | 7 (63.6%) | 10 (62.4%) |
| Male | 4 (40%) | 4 (36.3%) | 6 (37.5%) |
| Age, years (median, range) | 54 (46-68) | 61 (43-74) | 44 (31-62) |
| Phenotype<sup>1</sup> |  |  |  |
| ACM | 2 (20%) | 4 (36.3%) |  |
| DCM | 2 (20%) | 3 (27.3%) |  |
| HCM | 6 (60%) | 4 (36.3%) |  |
| Carrier status affected | 10 (100%) | 3 (27.3%) |  |
| Education level |  |  |  |
| Low | 4 (40%) | 2 (18.2%) |  |
| Medium | 1 (10%) | 4 (36.3%) |  |
| High | 5 (50%) | 5 (45.5%) |  |
| Family status |  |  |  |
| Single | 0 | 3 (27.3%) |  |
| With partner, no child | 2 (20%) | 0 |  |
| Alone, with child(ren) | 0 | 1 (9.1%) |  |
| With partner and child(ren) | 8 (80%) | 7 (63.6%) |  |
| Health literacy<sup>2</sup> |  |  |  |
| Low | 0 | 0 |  |
| Medium | 1 (10%) | 3 (27.3%) |  |
| High | 9 (90%) | 8 (72.7%) |  |
| Digital literacy<sup>2</sup> |  |  |  |
| Low | 1 (10%) | 1 (9.1%) |  |
| Medium | 0 | 4 (36.3%) |  |
| High | 9 (90%) | 6 (54.5%) |  |
| Profession |  |  |  |
| Clinical geneticist in training |  |  | 2 (12.5%) |
| Genetic counselor |  |  | 2 (12.5%) |
| Clinical geneticist |  |  | 7 (43.8%) |
| Laboratory specialist |  |  | 2 (12.5%) |
| Social worker |  |  | 3 (18.8%) |
| Experience in cardiogenetics, years<br>(median, range) |  |  | 12.0 (1-34) |

### Themes and subthemes

Five main themes were identified (Figure 1). The first theme involved the general attitude and perspectives of stakeholders towards acceptability of digital tools in cascade testing. The remaining themes were defined as ‘domains’ in which digital methods impact traditional, in-person clinical cascade testing: (2) family communication, (3) decision-making, (4) care relations, and (5) the genetic care system. Table 2 shows interview quotations to illustrate study findings.

**Figure 1.**
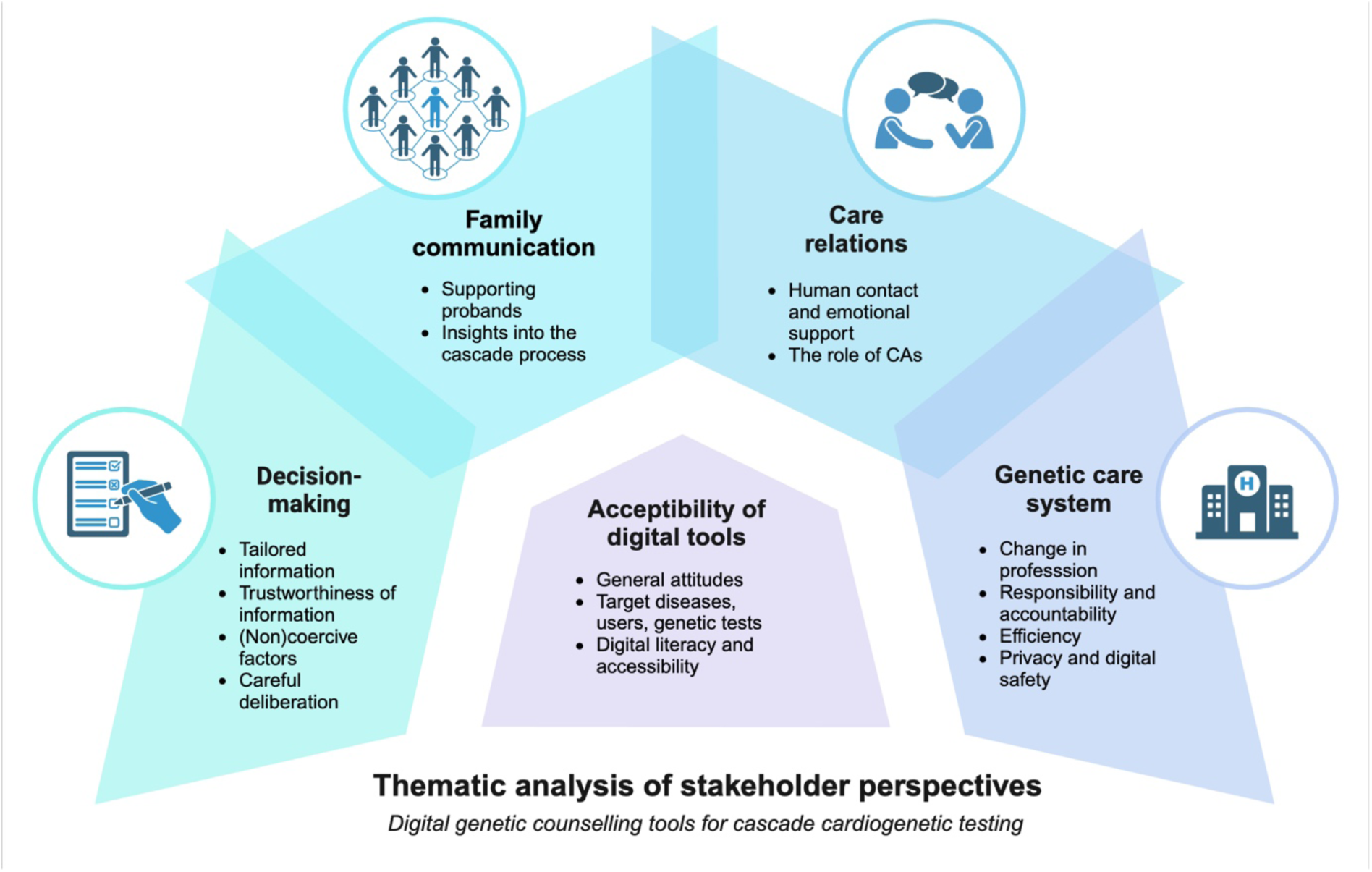
Result of thematic analysis of stakeholder perspectives on digital genetic counselling tools for cascade cardiogenetic testing. Schematic representation of themes (circles) and subthemes derived from thematic analysis of focus groups with probands, at-risk relatives and healthcare professionals. The central theme represents the acceptability of digital tools and the subthemes that were most relevant to acceptability according to participants. The outer circle represents the domains within regular cascade testing that would be impacted by digital tools, where subthemes capture the main opportunities and challenges identified by participants. CA= conversational agents, e.g. chatbot or virtual assistant.

**Table 2.**
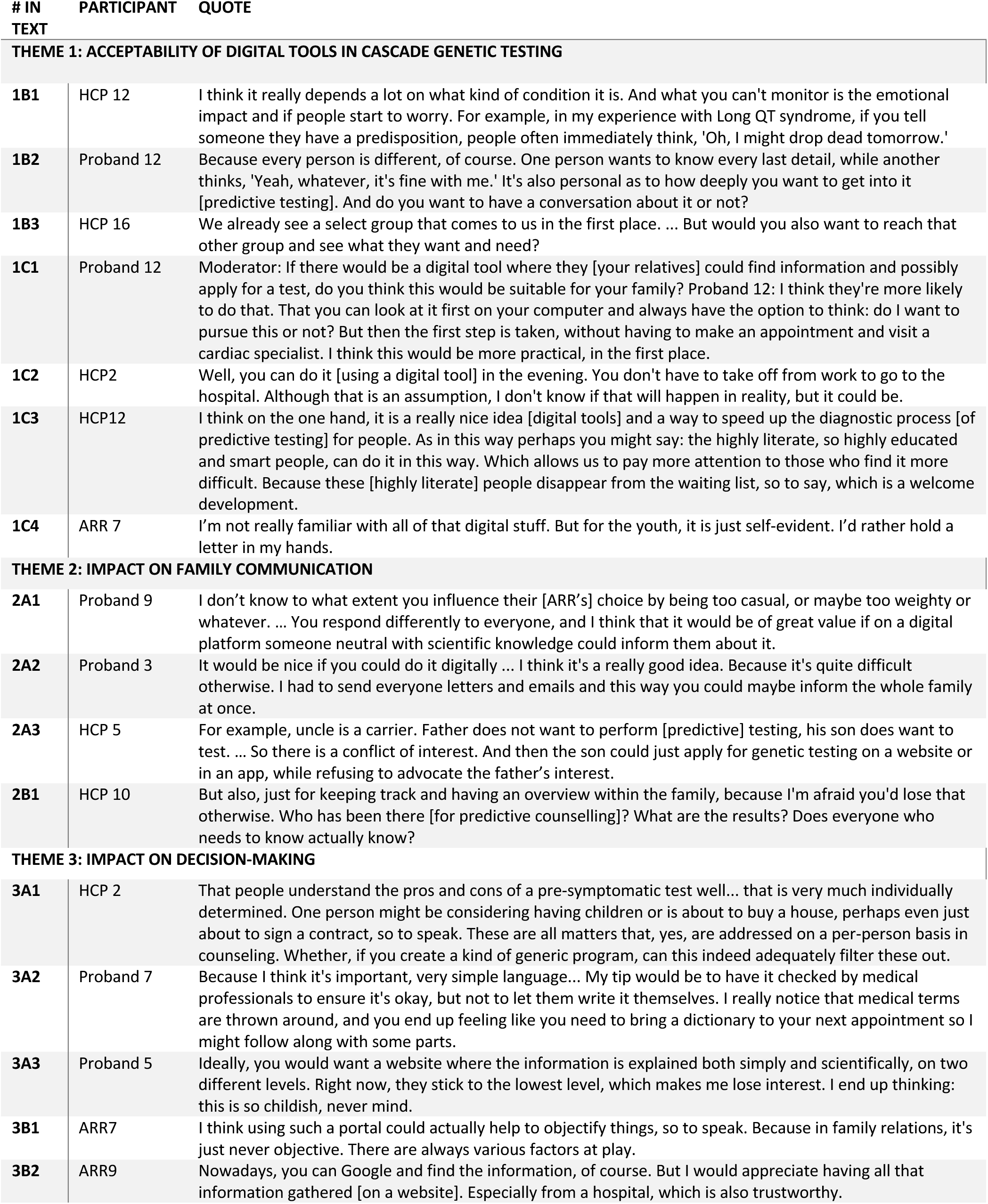
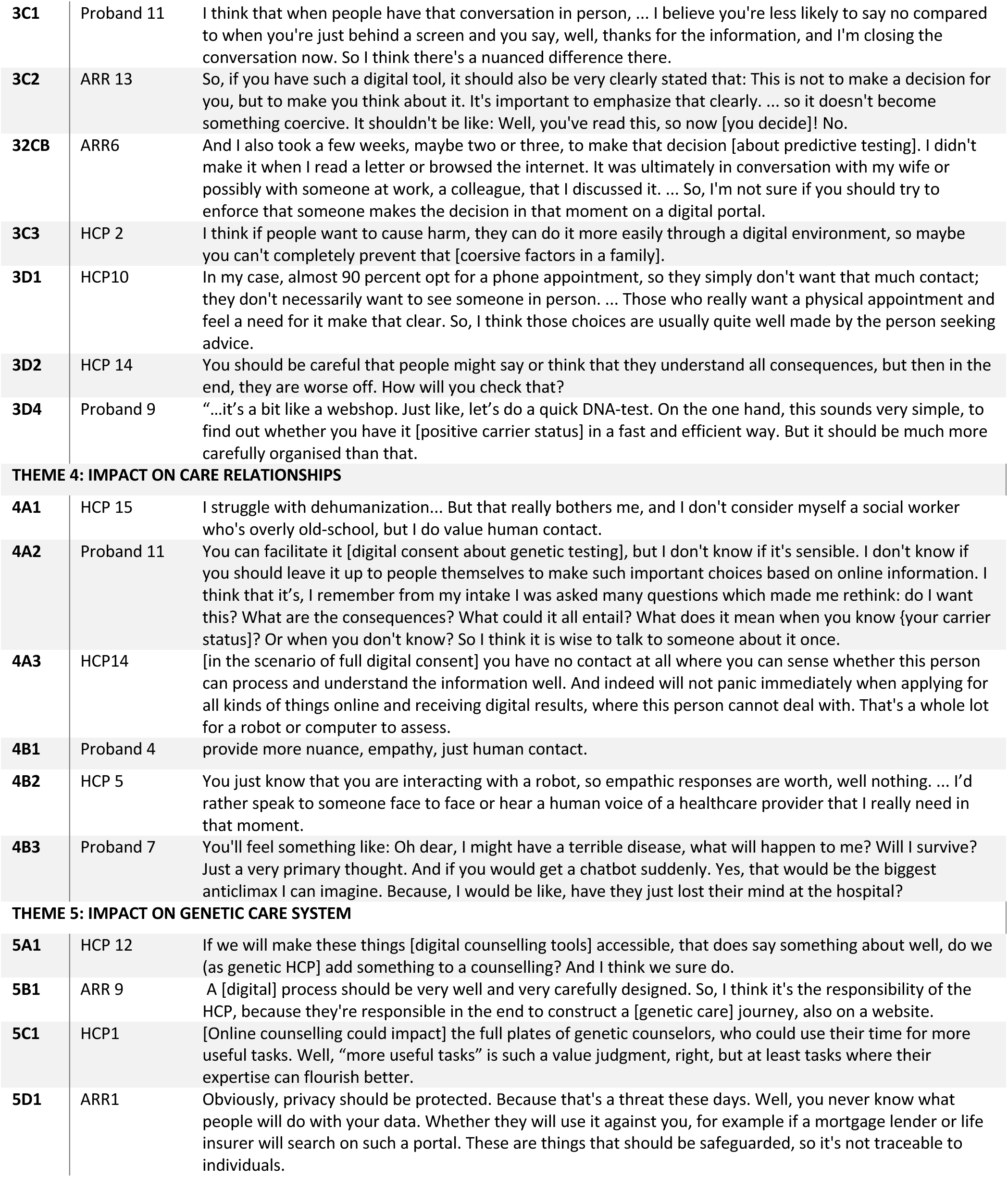
Participant quotations. Each quotation number corresponds with the related theme and subtheme of the quotation. Quotations are translated from Dutch. ARR= at-risk relative, HCP=healthcare professional.

| # IN TEXT | PARTICIPANT | QUOTE |
| --- | --- | --- |
| **THEME 1: ACCEPTABILITY OF DIGITAL TOOLS IN CASCADE GENETIC TESTING** |  |  |
| 1B1 | HCP 12 | I think it really depends a lot on what kind of condition it is. And what you can't monitor is the emotional impact and if people start to worry. For example, in my experience with Long QT syndrome, if you tell someone they have a predisposition, people often immediately think, 'Oh, I might drop dead tomorrow.' |
| 1B2 | Proband 12 | Because every person is different, of course. One person wants to know every last detail, while another thinks, 'Yeah, whatever, it's fine with me.' It's also personal as to how deeply you want to get into it [predictive testing]. And do you want to have a conversation about it or not? |
| 1B3 | HCP 16 | We already see a select group that comes to us in the first place. ... But would you also want to reach that other group and see what they want and need? |
| 1C1 | Proband 12 | Moderator: If there would be a digital tool where they [your relatives] could find information and possibly apply for a test, do you think this would be suitable for your family? Proband 12: I think they're more likely to do that. That you can look at it first on your computer and always have the option to think: do I want to pursue this or not? But then the first step is taken, without having to make an appointment and visit a cardiac specialist. I think this would be more practical, in the first place. |
| 1C2 | HCP2 | Well, you can do it [using a digital tool] in the evening. You don't have to take off from work to go to the hospital. Although that is an assumption, I don't know if that will happen in reality, but it could be. |
| 1C3 | HCP12 | I think on the one hand, it is a really nice idea [digital tools] and a way to speed up the diagnostic process [of predictive testing] for people. As in this way perhaps you might say: the highly literate, so highly educated and smart people, can do it in this way. Which allows us to pay more attention to those who find it more difficult. Because these [highly literate] people disappear from the waiting list, so to say, which is a welcome development. |
| 1C4 | ARR 7 | I'm not really familiar with all of that digital stuff. But for the youth, it is just self-evident. I'd rather hold a letter in my hands. |
| **THEME 2: IMPACT ON FAMILY COMMUNICATION** |  |  |
| 2A1 | Proband 9 | I don't know to what extent you influence their [ARR's] choice by being too casual, or maybe too weighty or whatever. ... You respond differently to everyone, and I think that it would be of great value if on a digital platform someone neutral with scientific knowledge could inform them about it. |
| 2A2 | Proband 3 | It would be nice if you could do it digitally ... I think it's a really good idea. Because it's quite difficult otherwise. I had to send everyone letters and emails and this way you could maybe inform the whole family at once. |
| 2A3 | HCP 5 | For example, uncle is a carrier. Father does not want to perform [predictive] testing, his son does want to test. ... So there is a conflict of interest. And then the son could just apply for genetic testing on a website or in an app, while refusing to advocate the father's interest. |
| 2B1 | HCP 10 | But also, just for keeping track and having an overview within the family, because I'm afraid you'd lose that otherwise. Who has been there [for predictive counselling]? What are the results? Does everyone who needs to know actually know? |
| **THEME 3: IMPACT ON DECISION-MAKING** |  |  |
| 3A1 | HCP 2 | That people understand the pros and cons of a pre-symptomatic test well... that is very much individually determined. One person might be considering having children or is about to buy a house, perhaps even just about to sign a contract, so to speak. These are all matters that, yes, are addressed on a per-person basis in counseling. Whether, if you create a kind of generic program, can this indeed adequately filter these out. |
| 3A2 | Proband 7 | Because I think it's important, very simple language... My tip would be to have it checked by medical professionals to ensure it's okay, but not to let them write it themselves. I really notice that medical terms are thrown around, and you end up feeling like you need to bring a dictionary to your next appointment so I might follow along with some parts. |
| 3A3 | Proband 5 | Ideally, you would want a website where the information is explained both simply and scientifically, on two different levels. Right now, they stick to the lowest level, which makes me lose interest. I end up thinking: this is so childish, never mind. |
| 3B1 | ARR7 | I think using such a portal could actually help to objectify things, so to speak. Because in family relations, it's just never objective. There are always various factors at play. |
| 3B2 | ARR9 | Nowadays, you can Google and find the information, of course. But I would appreciate having all that information gathered [on a website]. Especially from a hospital, which is also trustworthy. |
| 3C1 | Proband 11 | I think that when people have that conversation in person, ... I believe you're less likely to say no compared to when you're just behind a screen and you say, well, thanks for the information, and I'm closing the conversation now. So I think there's a nuanced difference there. |
| 3C2 | ARR 13 | So, if you have such a digital tool, it should also be very clearly stated that: This is not to make a decision for you, but to make you think about it. It's important to emphasize that clearly. ... so it doesn't become something coercive. It shouldn't be like: Well, you've read this, so now [you decide]! No. |
| 32CB | ARR6 | And I also took a few weeks, maybe two or three, to make that decision [about predictive testing]. I didn't make it when I read a letter or browsed the internet. It was ultimately in conversation with my wife or possibly with someone at work, a colleague, that I discussed it. ... So, I'm not sure if you should try to enforce that someone makes the decision in that moment on a digital portal. |
| 3C3 | HCP 2 | I think if people want to cause harm, they can do it more easily through a digital environment, so maybe you can't completely prevent that [coersive factors in a family]. |
| 3D1 | HCP10 | In my case, almost 90 percent opt for a phone appointment, so they simply don't want that much contact; they don't necessarily want to see someone in person. ... Those who really want a physical appointment and feel a need for it make that clear. So, I think those choices are usually quite well made by the person seeking advice. |
| 3D2 | HCP 14 | You should be careful that people might say or think that they understand all consequences, but then in the end, they are worse off. How will you check that? |
| 3D4 | Proband 9 | "...it's a bit like a webshop. Just like, let's do a quick DNA-test. On the one hand, this sounds very simple, to find out whether you have it [positive carrier status] in a fast and efficient way. But it should be much more carefully organised than that. |
| THEME 4: IMPACT ON CARE RELATIONSHIPS |  |  |
| 4A1 | HCP 15 | I struggle with dehumanization... But that really bothers me, and I don't consider myself a social worker who's overly old-school, but I do value human contact. |
| 4A2 | Proband 11 | You can facilitate it [digital consent about genetic testing], but I don't know if it's sensible. I don't know if you should leave it up to people themselves to make such important choices based on online information. I think that it's, I remember from my intake I was asked many questions which made me rethink: do I want this? What are the consequences? What could it all entail? What does it mean when you know {your carrier status}? Or when you don't know? So I think it is wise to talk to someone about it once. |
| 4A3 | HCP14 | [in the scenario of full digital consent] you have no contact at all where you can sense whether this person can process and understand the information well. And indeed will not panic immediately when applying for all kinds of things online and receiving digital results, where this person cannot deal with. That's a whole lot for a robot or computer to assess. |
| 4B1 | Proband 4 | provide more nuance, empathy, just human contact. |
| 4B2 | HCP 5 | You just know that you are interacting with a robot, so empathic responses are worth, well nothing. ... I'd rather speak to someone face to face or hear a human voice of a healthcare provider that I really need in that moment. |
| 4B3 | Proband 7 | You'll feel something like: Oh dear, I might have a terrible disease, what will happen to me? Will I survive? Just a very primary thought. And if you would get a chatbot suddenly. Yes, that would be the biggest anticlimax I can imagine. Because, I would be like, have they just lost their mind at the hospital? |
| THEME 5: IMPACT ON GENETIC CARE SYSTEM |  |  |
| 5A1 | HCP 12 | If we will make these things [digital counselling tools] accessible, that does say something about well, do we (as genetic HCP) add something to a counselling? And I think we sure do. |
| 5B1 | ARR 9 | A [digital] process should be very well and very carefully designed. So, I think it's the responsibility of the HCP, because they're responsible in the end to construct a [genetic care] journey, also on a website. |
| 5C1 | HCP1 | [Online counselling could impact] the full plates of genetic counselors, who could use their time for more useful tasks. Well, "more useful tasks" is such a value judgment, right, but at least tasks where their expertise can flourish better. |
| 5D1 | ARR1 | Obviously, privacy should be protected. Because that's a threat these days. Well, you never know what people will do with your data. Whether they will use it against you, for example if a mortgage lender or life insurer will search on such a portal. These are things that should be safeguarded, so it's not traceable to individuals. |

### Theme 1 – Acceptability of digital tools in cascade genetic testing

#### A. General attitudes

Stakeholders expressed a predominantly positive attitude towards digitisation of (parts of) family communication and cascade testing. If ARR or probands did not appreciate it for themselves, they envisioned digital pathways to be suitable for younger relatives. Benefits of digital tools included: improved accessibility, with reduced waiting times and the opportunity to relocate parts of genetic counselling to the private environment. However, stakeholders noticed general objections to digitisation of pre- or post-test counselling, or both, including lack of human contact, risking careful deliberation, and risk of decreasing quality of care standards.

#### B. Target diseases, users, and genetic tests

All stakeholder groups emphasised the importance of considering the target audience in designing and developing digital tools in three ways. First, some HCP stated that the acceptability of using digital predictive counseling tools depends on the characteristics of the targeted genetic condition, including disease severity, penetrance, and availability of treatment options. Clinically milder and treatable conditions, with moderate psychosocial impacts, were considered more suitable(Table 2; Q1B1). Many HCP argued that, for counselling cardiogenetic diseases specifically, much information is generic and therefore suitable for digital information transfer. For cascade testing, digital information provision was deemed appropriate for only specific (likely) pathogenic variants. For example, complexities in genetic counselling for variants of unknown significance (VUS) and some likely pathogenic variants might get lost in a digital context. HCP also worried about loss of insights into phenotype-specific characteristics in ARR and the family pedigree. This might result in a loss of tailored counselling of phenotypical characteristics in ARR that are not yet diagnosed.

Second, stakeholders argued that digital tool design should be adjusted to accommodate the needs of end users. Probands and HCP stated that the variety of ARR’s backgrounds complicates design, by illustrating why a “one size fits all” approach is not suitable for digital pathways for cascade testing(Q1B2). In their view, interactive modalities such as questionnaires or CA could, partly, accommodate varying needs. Some HCP suggested that digital tools should be specifically targeted towards groups that are currently underserved(Q1B3).

Finally, HCP recommended careful consideration in providing specific genetic tests to counselees in a digital setting. Some considered digital tools more appropriate for predictive genetic testing, compared to testing of symptomatic ARR. Few HCP argued that digital tools could contribute to diagnostic pre- and post-test counselling of probands, potentially in a mainstreaming setting. Others considered adopting digital tools in a diagnostic setting not suitable, expressing hesitation about such tools to counsel VUS, for example.

#### C. Digital literacy and accessibility

Participants across all stakeholder groups noted a potential lower threshold for ARR to access genetic counselling in a digital setting, as technology was considered widely accessible(Q1C1). Probands thought initiating genetic care would be easier in a digital setting, especially since the Dutch referral process through the GP using a family letter was considered complex. Also, the opportunity to access genetic counselling at a time and place convenient for the counselee was mentioned(Q1C2). Finally, one HCP noted that digital tools for cascade testing could potentially reach ARR who are who currently not receiving genetic counselling as they are reluctant to speak with a HCP or have a digital mindset.

However, most HCP argued that digital tools might disproportionally benefit already privileged counselees. They argued that families with strong digital and literacy skills, currently already overrepresented in Dutch genetics clinics, might get even greater access to genetic services. Some HCP stated that this may not necessarily be problematic, as shorter waiting lists create more opportunities for in-person counselling of less privileged counselees(Q1C3). All stakeholder groups agreed that digital counselling pathways are potentially less suitable for elderly with limited digital access and literacy(Q1C4). Most stakeholders felt that digital pathways would be most suitable for “younger” relatives, (i.e., those between adolescence and 50 years).

Several HCP emphasised that intuitive user design and stakeholder involvement are essential prerequisites for developing accessible digital genetic services. Stakeholders agreed that providers of digital tools should actively recognise digitally or health-illiterate counselees and offer support or alternative services.

### Theme 2 – Impact on family communication

#### A. Supporting probands in informing their relatives

ARR and probands both argued that current family communication in cascade testing relies heavily on the knowledge and efforts of probands. Many probands felt not equipped to address difficult questions from ARR while feeling responsible to inform them correctly and consistently(Table 2; Q2A1). One proband noted that ARR might feel hesitant to contact or ‘bother’ the proband for further information in an already emotionally stressful time. These stakeholders agreed that digital tools could lower the burden on probands, as ARR would have direct access to relevant information(Q2A2). Furthermore, stakeholders argued that digital tools could support the cascade process in situations with complex family relations, limited contact, or relatives living abroad. It might lower the threshold to reach out to ARR, either directly or through an HCP. However, HCP also expressed concerns that digital tools might limit options for HCP to intervene or support families with complex relations(Q2A3) due to limited involvement.

#### B. Insights into the cascade process

HCP believed digital tools could enhance insights into who is informed and engaged in cascade testing. However, they also considered it potentially challenging to integrate tools with electronic health records and to remain updated on cascade testing(Q2B1). Several HCP consider pedigrees as a relevant functionality to gain insight into family relations and inheritance. Similarly, some ARR believed pedigrees could also provide insights into carrier status and might motivate other ARR to perform cardiac screening or genetic testing.

### Theme 3 – Impact on decision-making

#### A. Tailored information in digital counselling tools

Many HCP emphasised the importance of tailoring digital pre-test counselling information to counselees’ life stage, personal values, and health literacy(Table 2; Q3A1). Most HCP expected this to be challenging in a digital context, while ARR and other HCP argued digital tools could improve information provision. For example, online information can be reread, unlike in-person conversations. Also, digital tools enable information transfer through various formats, such as text, videos, animations, or illustrations. Most probands and ARR emphasised that clear, concise, and basic information is essential for pre-test information provision, which was often not during in-person consultations(Q3A2). Other probands with high educational levels suggested a more layered approach, where complex information is available on demand(Q3A3).

#### B. Trustworthiness of information

ARR and probands considered a digital tool for informing ARR as more objective than information shared within a family, noting that family dynamics can influence how information is provided(Q3B1). Also, several ARR attempted to gather information through web search engines, which they often found frightening and confusing. In contrast, digital tools, hosted by established medical institutions, would be considered trustworthy and could assist ARR in finding relevant information for their families(Q3B2). Stakeholders across all groups agreed that digital counselling content should be reviewed by medical professionals.

#### C. (Non)coercive factors

One proband thought that digital pathways were less coercive towards genetic counselling or testing, since no HCP is present to advocate for a test(Q3C1). However, some ARR worried that a streamlined digital pathway might nudge an ARR towards quick decision-making. Several ARR explained that they needed time, ranging from days to weeks, to think, reflect, and discuss their decision. They argued that digital tools should explicitly allow time for reflection before consenting(Q3C2). Some ARR also believed that pursuing genetic testing is important, and felt that a digital platform should explicitly encourage ARR to undergo genetic testing or cardiac screening, or persuade those in doubt to do so(Q3C3). Additionally, some HCP emphasised that in a digital context they would have less insight into potential coercive influences within the family, such as relatives pressuring someone to make a specific decision, taking over the digital pathway, or to click for someone else(Q3C3). Ideally, these HCP wanted to filter out such cases but questioned the feasibility of doing so.

#### D. Careful deliberation

Several HCP stated that digital pathways might empower ARR in their decision-making and post-test processes, thereby promoting counselee-centred care. Based on their experience with current pathways, they found that counselees are often well-equipped to decide whether they prefer in-person, telephone, or video consultations(Q3D1). Digital pathways could serve as an additional option within genetic care if counselees always have the choice to opt-out of digital pathways.

Many HCP also shared concerns about the impact of digital tools on the quality of informed consent for genetic testing, especially without their involvement(Q3D2). Several participants, both HCP and probands, worried that digital pathways might oversimplify the decision-making process, reducing it to merely ticking a box. This was illustrated by the metaphor of an online webshop transaction, where ARR would opt for genetic testing without careful consideration(Q3D4). Most HCP believed that incorporating an in-person contact moment alongside a digital pathway is necessary to ensure ARR’s understanding. Some ARR and probands shared this view. In contrast, other HCP argued that for digitally skilled or already well-informed counselees, additional in-person counselling would not be required to obtain fully informed consent.

### Theme 4 – Impact on care relations

#### A. Human contact and emotional support

Stakeholders highly valued direct human contact between counselees and HCP in the current care. Some expressed concerns that further digitisation could dehumanise genetic care(Table 2; Q4A1). In-person contact, either in-person or via video/telephone, was often described as irreplaceable, especially for decision-making and psychosocial support(Q4A2). While direct human contact was not deemed essential for providing standard information or collecting medical and family history, it was considered crucial for adequately interpreting genetic results by both HCP and patients and for clinical follow-up by HCP(Q4A3). Additionally, a few HCP highlighted the importance of building a care relation during pre-test consultation to prepare for the potential return of positive test results.

Several HCP noted that digital tools could allow for more meaningful contact for those requiring additional attention. Several stakeholders believed that for many ARR digital counselling might be a valuable contribution, under the condition that in-person counselling remains an option. Few HCP perceived mandatory in-person contact moments as paternalistic.

#### B. The role of conversational agents in genetic counselling

Several ARR and probands argued that digital alternatives for in-person contact, such as CA or direct chat functions with HCP, would not be adequate substitutes(Q4B1). Stakeholders also believed that developers should be cautious when developing CA for psychosocial support. Several HCP emphasised that only HCP can provide the necessary empathy and nuance(Q4B2). Some ARR and probands expressed agitation and felt upset when visualising ‘empathetic’ responses of CA(Q4B3). Some of them even expected counselees to disengage from digital counselling if confronted with a CA that fails to provide satisfactory answers or comes across as overly empathetic. Less emotionally charged topics or information were considered suitable for being transferred via CA if well-tailored to end-users. Some HCP stated they were unaware of technological capabilities of CA in the (near) future and whether they could provide thoughtful or empathetic answers. If so, their stance on implementing CA might change.

### Theme 5 – Impact on the genetic care system

#### A. Changes in the profession of genetic counsellors

Some HCP expressed concerns that transferring counselling tasks to digital tools could alter the profession of HCP in clinical genetic care. Several HCP described a potential shift in professional identity; in what it means to be a genetic HCP. One HCP worried that the existence of digital counselling tools might question the added value of HCP in genetic care, especially in the absence of human contact(Q5A1). Additionally, several HCP felt that digital counselling tools could diminish their job satisfaction.

#### B. Responsibility and accountability for digital counselling tools

To ensure the quality of a (partial) digital genetic care pathway, probands and ARR agreed that HCP must be responsible for both the quality of the information that is provided and the individual counselling process(Q5B1). Therefore, HCP should be closely involved in the design and development of digital counselling tools. While many HCP underlined these considerations, some argued that in digital pathways without HCP involvement, the responsibility should rest with the clinical centre providing the digital genetic service. If individual HCP are responsible for fully digital counselling pathways, they should have control over the information provided, especially when genetic test results are returned digitally. Also, it was argued that HCP should be linked to individual counselees to answer questions accurately, and ensure procedural quality. A few HCP highlighted the responsibilities of technical developers in maintaining quality of online genetic care pathways. HCP argued that implementing digital tools would delegate tasks to ARR of identifying misunderstandings or concerns and to reach out to HCP accordingly. While many HCP questioned whether it is appropriate to place these responsibilities on counselees, some felt it would be for most ARR.

#### C. Efficiency

All stakeholders perceived absence of travel times as an advantage of digital tools. Some probands noted that digital methods to inform ARR could save time compared to in-person conversations, especially with more distant ARR. Most HCP anticipated that digital tools would drastically reduce counselling time, allowing room for more complex consultations or tasks(Q5C1). One HCP was more sceptical, arguing that predictive counselling consultations are already relatively short and administrative tasks demand a lot of time. Several HCP expected digital tools to reduce waiting lists for genetic counselling. Cost impact was mentioned as another, albeit less emphasised, opportunity: some ARR and HCP expected lower healthcare costs, compared to traditional counselling.

#### D. Privacy and digital safety

To ensure responsible implementation, all stakeholder groups flagged the importance of digital safety and privacy. Not only to promote trust but also to safeguard medical and genetic information. Several ARR stressed that insurance companies should not have access to digital cascade testing tools(Q5D1). Several stakeholders expressed hesitance about using digital tools due to privacy concerns. To address these, secure login methods were suggested along with ensuring anonymity of users. Furthermore, data should not be shared within families without explicit consent.

## Discussion

This focus group study revealed a predominantly positive attitude among probands, ARR, and HCP towards digitisation of certain aspects of the cascade testing journey including predictive counselling. Expected benefits include saving in-person counselling time while increasing time for ARR to engage with relevant information, improving information provision at home, and increased accessibility of genetic counselling. However, specific concerns were raised on the applicability of CA for emotional support and the lack of in-person contact when implementing a fully digitised cascade testing pathway. Furthermore, to promote equitable genetic care, digital genetic testing tools should accommodate counselees with varying levels of (digital) literacy. This aligns with a recent study where HCP emphasised the need to prevent digital tools from further increasing existing inequalities in access to genetic care [29]. Accessibility for digitally illiterate counselees was already identified as one of the main challenges of implementing BRCA-DIRECT, a digital germline genetic testing pathway for breast cancer patients [23]. Our findings suggest that providers of digital tools have an active duty to recognise digitally or health-illiterate counselees and offer support or alternative services.

Stakeholders in this study noted that digital tools can both support and challenge sound decision-making. Safeguarding access to human contact during digital counselling was the key requirement for clinical integration. Without in-person HCP involvement, they worried that digital counselling methods would not ensure personalised decision-making. *Shickh et al.* already showed that in-person consultations following digital pre-test counselling enhanced personalised decision-making aligning with personal values and informed dialogue, compared to in-person counselling alone [30]. This substantiates that digital tools can benefit decision-making. Further research is needed to assess potential decisional conflict in counselees, and to understand the impact of fully digitised counselling pathways on decision-making and quality of informed consent.

There is limited knowledge about which parts of the genetic care pathway are suitable for digitisation. Recently, *Lee et al.* found that HCP were more positive about adopting digital tools in the pre-test rather than the post-test phase [29]. This partly aligns with our findings, though our stakeholders, including probands and ARR, were more sceptical about fully digitising pre-test counselling and decision-making for predictive testing. Similarly, HCP in *Lee et al.’s* study favoured hybrid models (part in-person, part digital counselling) for complex cases, while digital tools alone were deemed sufficient for less complex cases [29]. However, most stakeholders in our study considered a “human in the loop” necessary in all cases. This contrast may be attributed to participants’ tendency of hesitance towards digitisation, whereas *Lee et al.’s* cohort was described as prone towards digitisation [29]. Also, the age of ARR in our study was relatively high. All stakeholders expected young adult ARR to be even more positive about digital tools. A chatbot evaluation study by *Schmidlen et al.* underlined this view in relatives [20].

Participants’ critical attitude towards CA for psychosocial support aligns with a study by *Luca et al.* [31] where HCP considered complex tasks like explaining results or providing emotional support as unsuitable for CA. Instead, implementing digital tools may allow HCP to have more meaningful interactions with other counselees. An evaluation study of the Rosa chatbot, which provides factual information about BRCA testing to breast- and ovarian cancer patients alongside genetic counselling, found high patient satisfaction and trust [26]. However, participants did miss eye contact and human empathy while using the chatbot [26]. Both studies emphasise that CA in genetic services should augment rather than fully replace in-person contact [26, 31].

Strengths of our study include in-depth perspectives from representative stakeholders in cardiogenetics. Incorporating views from probands, ARR, and HCP offer unique insights into the perspectives and preferences within these groups. Although the focus is on cardiogenetic counselling, the themes and subthemes described are broadly applicable to other diseases and areas within (predictive) genetic counselling. A potential limitation of this study is selection bias. ARR who did not receive counselling were unfortunately not represented, resulting in an underrepresentation of perspectives of those hesitant about genetic testing. Furthermore, all participants expressed support for genetic testing, which might have influenced the perspectives shared as they were motivated to support methods that might increase cascade testing uptake.

In conclusion, digital tools for cardiogenetic counselling are promising in supporting both counselees and HCP in cascade testing. In the clinical setting of predictive counselling, efforts should focus on ensuring access to genetic services for all ARR and protecting quality of care and in-person involvement of HCP. Further research is necessary to enhance understanding of the opportunities and concerns identified in this study, both conceptually and empirically.

## Data availability

Anonymised data are available from the corresponding author upon reasonable request.

## Code availability

Not applicable.

## Funding

This project is financially supported by ZonMW/IMDI and the Dutch Heart Foundation. Grant number 104021006 (ZonMW) 2019B012 (Dutch Heart Foundation).

## Acknowledgements

We thank M. Hendriks and I. Dröge for contributing to the practical organisation and observation during the focus groups. We thank L. Yeates for her contribution to the final manuscript. Figure 1 (https://BioRender.com/f15w704) and the graphical abstract (https://BioRender.com/d23e144) were created in BioRender. van Tintelen, P. (2025).

## Author Contribution Statement

Conceptualisation: MvL, LvdH, PvT, NG; Funding acquisition LvdH, PvT, DH; Organisation: MvL; Data collection: MvL, NG, LvdH; Data analysis: MvL, SvT, LvdH; Writing original draft: MvL; Review and editing manuscript: MvL, SvT, LvdH, PvT, RK, TB, DH, MA, MC; Supervision: LvdH, PvT.

## Ethical approval

The study protocol was exempted from approval by the Medical Ethical Committee NedMec, because the Act of Medical Research Involving Human Subjects (WMO) was not applicable (research proposal no. 21-474/C). We obtained written and verbal informed consent of all participants in the study.

## Competing Interests

Authors declare no conflicts of interest.

## Notes

### Competing Interest Statement

The authors have declared no competing interest.

### Funding Statement

This study was funded by ZonMW/IMDI and the Dutch Heart Foundation. Grant number 104021006 (ZonMW) 2019B012 (Dutch Heart Foundation).

### Author Declarations

Medical Ethical Committee NedMec waived ethical approval for this work, because the Act of Medical Research Involving Human Subjects (WMO) was not applicable (research proposal no. 21-474/C).

